# COVID-19 and Influenza Infection Prevention and Control Measures in U.S. Jails, Prisons, and Detention Centers: A Scoping Review

**DOI:** 10.1101/2023.10.04.23296518

**Authors:** Leo Knudsen Westgard, Yvane Ngassa, Emily Grussing, Alysse G. Wurcel, Max Jordan Nguemeni Tiako

**Affiliations:** Division of Geographic Medicine and Infectious Diseases, Department of Internal Medicine, Tufts Medical Center; Tufts University School of Medicine; Department of Internal Medicine, Brigham and Women’s Hospital; Harvard Medical School

**Author notes:** **Corresponding Author:** Name: Alysse Wurcel.

**Keywords:** Jails, Prisons, Detention, Incarceration, Corrections, Influenza, COVID-19, SARS-CoV-2, Infection Prevention, Outbreak Management, Pandemic Preparedness

## Abstract

**Background:** People who are incarcerated or detained are at a high risk of infection and death due to viruses spread through droplets, including SARS-CoV-2 (COVID-19) and influenza. Increased risk is largely associated with structural and institutional conditions of detainment, including but not limited to congregate settings, poor ventilation, and barriers to accessing care. The lessons learned from managing previous infectious outbreaks can inform preventative measures for future emerging outbreaks.

**Aims:** Our project aims to analyze infection prevention and control measures for COVID-19 and influenza in U.S. correctional facilities.

**Methods:** We searched the PubMed and Embase databases for manuscripts evaluating the effectiveness of influenza or COVID-19 mitigation measures. Then, we reviewed the reference lists from our results to identify manuscripts not captured by the initial search.

**Results:** Out of the 553 articles initially found, only 28 met the inclusion criteria, with 27 focusing on COVID-19. Two additional studies were added after reviewing reference lists. Effective measures to prevent the spread of both infections included vaccines, testing, de-densification, limiting movement, masks, and contact tracing.

**Conclusion:** Improved infrastructure, stakeholder collaboration, and research on sustainable responses are needed to address disproportionate impacts on people who are incarcerated or detained.

## Introduction

Criminal-legal involved populations, including people who are detained and incarcerated in jails, prisons, and detention centers, are more likely to be infected with and die from viral respiratory infections. COVID-19 shined a spotlight on the structural conditions of carceral settings that facilitate the transmission of respiratory pathogens: overcrowding, a high number of visitors, poor ventilation, and inadequate access to soap, water, and personal protective equipment (Beaudry et al., 2020; Bick, 2007; Hammett et al., 2002; LeMasters et al., 2022; Liu et al., 2022; Montoya-Barthelemy et al., 2020; Norton & Heiss, 2020; Reinhart & Chen, 2020). The synergistic impact of an aging incarcerated population, higher rates of chronic diseases, including substance use disorder, and suboptimal access to healthcare in carceral settings leads to worse outcomes from COVID-19 (Dir et al., 2022; Hawks et al., 2020; Saloner et al., 2020; Strodel et al., 2021).

The spread of respiratory infectious diseases in detention facilities is a public health crisis that tracks back to the earliest days of using jails and prisons for public safety. The first published report of infectious diseases in jails is from 1896 and focused on tuberculosis (Chaucer, 1955). With increased awareness of tuberculosis mitigation, the focus of most public health publications on jails and prisons has been on two respiratory infections: influenza and COVID-19. Influenza outbreaks had very high attack rates in U.S. jails and prisons during the 1918 H1N1 virus (Kolata, 1999), 1957-1958 H2N2 virus (Cobos et al., 2016), 1968 H3N2 virus (Jester et al., 2020), and 2009 H1N1pdm09 virus (Reutter, 2010). Previous studies have reviewed literature on the health impacts of COVID-19 in carceral settings (Kim et al., 2022) and outbreak management strategies for highly contagious diseases in prisons (Beaudry et al., 2020). A previously published systematic review of COVID-19 prevention strategies highlighted a dearth of data on vaccination and quarantine (Esposito et al., 2022). This systematic review limited data collection to COVID-19 and publications before November 2021, leaving room for an updated analysis that incorporates studies reporting on mitigation in jails and prisons through the end of 2022.

There has been increased momentum to broaden pandemic preparedness plans past COVID-19 and learn from other respiratory infections. Although there have been strides in developing and operationalizing COVID-19 public health measures, including broad access to effective vaccination, COVID-19 continues to mutate, and public health authorities continue to express concern about future waves of transmission with associated increased morbidity and mortality. Outside of the COVID-19 pandemic, with continued climate change, experts expect an increasing frequency of new infections with potentially severe consequences for people residing in congregate settings (Carlson et al., 2022; Vargas-Parada, 2022). Literature on influenza and COVID-19 has not been evaluated as a joint entity, despite both infections sharing non-pharmacological interventions (Shi et al., 2022). Recognizing gaps in the literature from learning collectively from influenza and COVID-19, we conducted a scoping review of the effectiveness of influenza and COVID-19 mitigation strategies with the goal of developing recommendations for pandemic preparedness in jails and prisons.

## Methods

Three of the authors (AGW, MT, and YN) collaborated on a previous analysis (Tiako et al., 2022), which provided the scaffold for this study. The current study follows PRISMA reporting guidelines (Tricco et al., 2018) and the framework established by Arksey and O’Malley, but excludes the optional consultation exercise (Arksey & O’Malley, 2005). The final research question is, “What methods effectively mitigate the spread of influenza and COVID-19 outbreaks in U.S. prisons, jails, and detention centers?”

### Search Strategy

The search was conducted in November and December of 2022 using the MEDLINE/PubMed and Embase databases. Final terms, developed in consultation with a Tufts University School of Medicine librarian, included: COVID-19, SARS-CoV-2, influenza, flu, prison, jail, carceral, detention, detain, incarcerate, correctional, and penal (Appendix 1). Results were restricted to U.S. studies with the MeSH term, “United States.” All results not reporting real-world data on the effectiveness of an intervention on incarcerated or detained were excluded. Articles published in languages other than English and articles published before 1982 were also excluded using PubMed filters. Articles outside of the United States were excluded because of interest in informing domestic policy decisions.

### Study Selection and Screening

The results were independently screened by two reviewers (LKW and YN) using a three-stage process. First, to assess the relevance, both reviewers screened results by title and then abstract. Second, the full texts of selected articles were reviewed. Both reviewers met throughout the process to ensure inter-rater reliability between reviewers and to resolve any conflicts. Third, references from all results were manually scanned at the end of the process to find any results not captured by the initial search. Any relevant manuscripts revealed through step three were then run through steps one and two.

### Data Abstraction

Two authors (LKW and YN) independently extracted data from selected articles. Data were collected on the title, author, infection, intervention type, study design, whether a study was focused on prevention or outbreak management, population size, study setting, geographic location, and the results of each study. Microsoft Excel Version 2211 was used to chart and summarize data.

## Results

The original search yielded 250 PubMed/MEDLINE results and 329 Embase results. 553 articles were excluded, including 143 duplicates, leaving 26 results meeting the inclusion criteria (Figure 1). Following the screening of references, we included two additional manuscripts that met the eligibility criteria but did not appear in our initial search, meaning that there were 28 total manuscripts. 27 results discussed interventions related to COVID-19, while only one discussed influenza (Table 1). Nearly all results for both infection types were observational, with most analyzing cross-sectional data. No results discussed mitigation strategies in ICE detention centers, and only two results discussed them in youth detention centers (Goldman et al., 2022; Unruh et al., 2021). We will discuss the results in 6 subsections: vaccines, testing, de-densification, limited movement, masks, and contact tracing.

**Figure 1.**
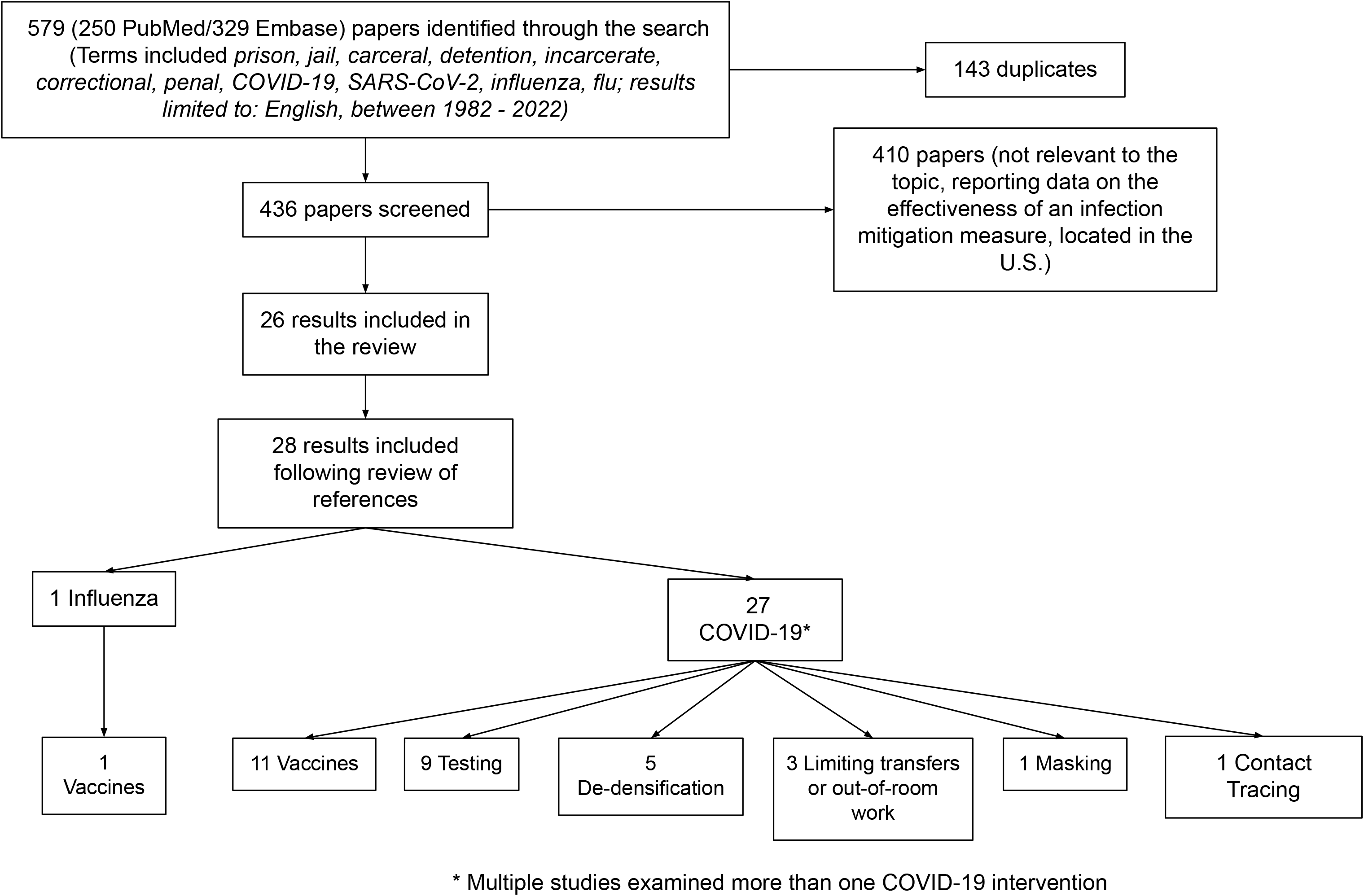

**Table 1.**

| Title | Author | Disease/<br>Infection | Topic/Intervention Type | Study type | Pandemic<br>preparedness<br>(0) or<br>emergency<br>response (1) | Sample size | Setting (Jail,<br>Prison, Detention<br>center) | Location | Findings |
| --- | --- | --- | --- | --- | --- | --- | --- | --- | --- |
| The utility of video technology and enhanced infection control in reducing COVID-19 disease burden in a custodial setting | Unruh | COVID-19 | Contact tracing using video-monitoring | Cohort | 1 | 505 | Juvenile detention center | IL | Contact tracing and prompt diagnosis by an outbreak management team reduced attack and positivity rates |
| Prison Population Reductions and COVID-19: A Latent Profile Analysis Synthesizing Recent Evidence From the Texas State Prison System | Vest | COVID-19 | Decarceration (reducing housing capacity) | Ecological, longitudinal | 1 | 103 Texas Prisons | Prisons and jails | TX | Reduced housing capacity was associated with lower risk of infection and death |
| Association Between Prison Crowding and COVID-19 Incidence Rates in Massachusetts Prisons, April 2020-January 2021 | Leibowitz | COVID-19 | Decarceration + limiting overcrowding | Ecological, longitudinal | 1 | 6,876 | Prisons | MA | Decreased prison density was associated with decreased incidence |
| Epidemiology of COVID-19 Among Incarcerated Individuals and Staff in Massachusetts Jails and Prisons | Jimenez | COVID-19 | Decarceration, Asymptomatic testing | cohort, retrospective observational | 1 | 14,987 | Both | MA | Incidence was lower in facilities that released more of their baseline population. Asymptomatic testing was effective in identifying cases |
| COVID-19 in the California State Prison System: an Observational Study of Decarceration, Ongoing Risks, and Risk Factors | Chin | COVID-19 | Decarceration, reduced labor participation | Cross-sectional | 1 | 119,401 | Prisons | CA | Dormitory style housing and labor participation was associated with higher incidence |
| The association between intersystem prison transfers and COVID-19 incidence in a state prison system | Brinkley-Rubinstein | COVID-19 | Limiting movement of incarcerated people in/out of prisons (limiting transfers) | Longitudinal | 1 | > 20,000 transfers | Prisons | CA | Transfers between prisons was associated with higher future case rates |
| COVID-19 Outbreaks in Correctional Facilities with Work-Release Programs — Idaho, July–November 2020 | Dunne | COVID-19 | Limiting movement of incarcerated people in/out of prisons (suspension of work-release programs) | Longitudinal | 1 | 760 | Unspecified | ID | Suspension of work-release programs may lead to decreased incidence |
| COVID-19 Preventive Measures in Northern California Jails: Perceived Deficiencies, Barriers, and Unintended Harms | Liu | COVID-19 | Masking, implications for de-densifying housing, decarceration | Cross-sectional: mixed methods | 1 | 788 jail residents and 380 staff | Jails | CA | Smaller cell size relative to larger cell size and weekly access to new masks were associated with reduced incidence |
| Serial Laboratory Testing for SARS-CoV-2 Infection Among Incarcerated and Detained Persons in a Correctional and Detention Facility — Louisiana, April–May 2020 | Njuguna | COVID-19 | Testing | Cross-sectional | 1 | 98 | Unspecified | LA | Testing close contacts of COVID-19-positive individuals was effective in identifying cases that were not identified through symptom screening |
| Outbreak of COVID-19 and interventions in a large jail — Cook County, IL, United States, 2020 | Zawitz | COVID-19 | Testing | Longitudinal | 1 | 628 | Jail | Cook County, IL | Asymptomatic testing identified a high number of cases |
| Aiming for Zero: Reducing Transmission of Coronavirus Disease 2019 in the D.C. Department of Corrections | Epting | COVID-19 | Testing | Cohort | 1 | 1,330 | Jail | D.C. | Hot spot testing is an effective strategy for identifying symptomatic and asymptomatic cases |
| COVID-19 in the New York City Jail System: Epidemiology and Health Care Response, March–April 2020 | Chan | COVID-19 | Testing | Cohort, retrospective observational | 1 | 978 | jails | NY, NY | Both symptomatic and asymptomatic testing was effective in identifying a high number of cases |
| Identification of Presymptomatic and Asymptomatic Cases Using Cohort-Based Testing Approaches at a Large Correctional Facility—Chicago, Illinois, USA, May 2020 | Wadhwa | COVID-19 | Testing (cohort-based) | Quasi-experimental | 1 | 224 | Jail | Cook County, IL | Serial and asymptomatic testing was effective in identifying a high number of cases |
| Correctional-Facility-Outbreak-Associated COVID-19 Cases Among Asymptomatic Persons Identified Through Universal Testing: Vermont, 2020 | Pringle | COVID-19 | Testing (facility-wide) | Cross-sectional | 1 | 197 residents and 115 staff | Unspecified | VT | Broad-based testing was more effective at identifying cases than symptom screening |
| Mass Testing for SARS-CoV-2 in 16 Prisons and Jails — Six Jurisdictions, United States, April–May 2020 | Hagan | COVID-19 | Testing (mass) | Cross-sectional | 1 | 41,454 | Prisons and jails | Various | Mass testing was significantly more effective at identifying cases than symptom-based testing |
| Mass SARS-CoV-2 Testing in a Dormitory-Style Correctional Facility in Arkansas | Tompkins | COVID-19 | Testing (mass, asymptomatic included) | Cross-sectional | 1 | 1,647 | Unspecified | AR | Asymptomatic testing limits virus transmission |
| Rapid Transmission of Severe Acute Respiratory Syndrome Coronavirus 2 in Detention Facility, Louisiana, USA, May-June, 2020 | Wallace | COVID-19 | Testing (serial) | Cross-sectional | 1 | 143 | Jail | LA | Serial and asymptomatic testing may help interrupt transmission |
| Breakthrough SARS-CoV-2 Infections in Prison after Vaccination | Brinkley-Rubinstein | COVID-19 | Vaccination | Retrospective cohort | 1 | 2,380 (including staff) | Both | RI | Vaccination of staff and incarcerated persons likely prevents virus transmission |
| Strategies for Addressing Vaccine Hesitancy Within California State Prisons in 2021 and Beyond | Garcia-Grossman | COVID-19 | Vaccination outreach | Cohort | 1 | Unspecified—less than 5,500 | Prisons | CA | A vaccine education initiative effectively improved vaccination rates at two different jails by 10% and 19% respectively |
| Effectiveness of the mRNA-1273 Vaccine during a SARS-CoV-2 Delta Outbreak in a Prison | Chin | COVID-19 | Vaccination | Cohort | 1 | 827 | Prison | CA | Vaccination was quite effective against a SARS-CoV-2 delta outbreak |
| Vaccine Effectiveness during Outbreak of COVID-19 Alpha (B.1.1.7) Variant in Men's Correctional Facility, United States | Silverman | COVID-19 | Vaccination | Cohort | 1 | 854 | Prison | VA | Vaccination was highly effective at preventing infection |
| Effectiveness of Coronavirus Disease 2019 (COVID-19) Vaccines Among Incarcerated People in California State Prisons: Retrospective Cohort Study | Chin | COVID-19 | Vaccination | Cohort (retrospective) | 1 | 60,707 | Prisons | CA | mRNA vaccines were highly effective at preventing infection |
| Outbreak of SARS-CoV-2 B.1.617.2 (Delta) Variant Infections Among Incarcerated Persons in a Federal Prison - Texas, July-August 2021 | Hagan | COVID-19 | Vaccination | Cross-sectional case study (retrospective) | 1 | 233 | Prison (federal) | TX | Vaccinated individuals were less likely to become infected with, become hospitalized, or die than vaccinated individuals |
| Infectiousness of SARS-CoV-2 breakthrough infections and reinfections during the Omicron wave | Tan | COVID-19 | Vaccination | Cohort | 1 | 1,226 | Prisons | CA | Vaccination including the receipt of booster vaccines reduced the transmissibility of the virus |
| Protection against Omicron conferred by mRNA primary vaccine series, boosters, and prior infection | Chin | COVID-19 | Vaccination | Case control | 1 | 195,952 | Prisons | CA | A third dose of an mRNA vaccine provided additional protection over two doses. |
| Waning of Vaccine-Conferred Protection against SARS-CoV-2 Infection: Matched Case-Control Test-Negative Design Study in Two High-Risk Populations | Goldhaber-Fiebert | COVID-19 | Vaccination | Case control | 1 | 25,970 | Prisons | CA | Vaccination is highly effective in preventing infection but effectiveness wanes over time |
| Vaccination for Justice-Involved Youth | Goldman | COVID-19 | Vaccination outreach | Case study (self-identified) | 1 | 108 | Juvenile detention centers | Allegheny County, PA | Individualized education and counseling along with community-based vaccine clinics were effective in vaccinating justice-involved youth |
| Distribution of A(H1N1)pdm09 Influenza Vaccine: Need for Greater Consideration of Smaller Jails | Lee | Flu | Vaccination (pandemic response planning) | Cross sectional, mixed-methods analysis | 0 | 447 facilities | Jails | U.S. | Having a pandemic response plan was associated with a greater likeliness of jails receiving H1N1 vaccines to distribute |

### Mitigation Strategy 1: Vaccines

Lee et al. (Lee et al., 2014) modeled predictors of administering influenza vaccines by conducting a cross-sectional survey with qualitative and quantitative sections at over 1,000 correctional facilities during the 2009 H1N1 pandemic. Compared with jails without a pandemic plan, jails that reported having a pandemic plan were 69% more likely to obtain the influenza vaccine to distribute to residents (95% CI; 1.19, 1.45) (Lee et al., 2014). Ten studies were on COVID-19 vaccination in detention facilities (Brinkley-Rubinstein, Peterson, et al., 2021; Chin, Leidner, et al., 2021; Chin, Leidner, Lamson, et al., 2022; Chin, Leidner, Zhang, et al., 2022; Garcia-Grossman et al., 2022; Goldhaber-Fiebert et al., 2022; Goldman et al., 2022; Hagan et al., 2021; Silverman et al., 2022; Tan et al., 2023). Silverman et al. (Silverman et al., 2022), Chin et al. (Chin, Leidner, et al., 2021), Chin et al. (Chin, Leidner, Zhang, et al., 2022), Brinkley-Rubinstein et al. (Brinkley-Rubinstein, Peterson, et al., 2021), and Tan et al. (Tan et al., 2023) used cohort study designs to demonstrate that vaccination is an effective strategy for COVID-19 prevention in prisons. When reported, effectiveness of vaccination against infection ranged between 56.6% (Chin, Leidner, et al., 2021) and 97% (Chin, Leidner, Zhang, et al., 2022). Tan et al. (Tan et al., 2023) showed that vaccination without prior infection reduced an index case’s risk of transmitting infection by 22% (CI, 6-36%). Using retrospective cross-sectional study designs, Hagan et al. (Hagan et al., 2021) and Brinkley-Rubinstein et al. (Brinkley-Rubinstein, Peterson, et al., 2021) showed the infection rate within a detained population was significantly higher for unvaccinated individuals compared with vaccinated individuals. Two studies, Chin et al. (Chin, Leidner, Lamson, et al., 2022) and Goldhaber-Fiebert et al. (Goldhaber-Fiebert et al., 2022), highlighted the need for booster doses of mRNA vaccines in California state prisons using case-control study designs. Goldman et al. (Goldman et al., 2022) studied a multi-faceted outreach initiative at a juvenile detention center with a virtual forum, one-on-one outreach to guardians, motivational interviewing, and coordination with community leaders. While this program focused on youths residing at the detention center, individuals in the county’s Community Intensive Supervision Program were also eligible. Out of 56 people receiving counseling through this program, 31 were vaccinated because of the initiative (Goldman et al., 2022). Garcia-Grossman et al. (Garcia-Grossman et al., 2022) evaluated the impact of vaccination education events with informal question and answer sessions, trivia, and incentives for participation. At these events, 10% of unvaccinated detainees in one prison and 19% in another prison received their first COVID-19 vaccine.

### Mitigation Strategy 2: Increased Testing

No studies evaluated the role of influenza testing as a mitigation strategy. Ten results assessed the use of COVID-19 testing in detention facilities (Chan et al., 2021; Epting et al., 2021; Hagan et al., 2020; Jimenez et al., 2020; Njuguna et al., 2020; Pringle et al., 2022; Tompkins et al., 2021; Wadhwa et al., 2021; Wallace et al., 2021; Zawitz et al., 2021). All results highlighted testing as an important strategy to interrupt the transmission of COVID-19 because testing identifies a high number of cases that would not be identified through symptom screening alone (Chan et al., 2021; Epting et al., 2021; Hagan et al., 2020; Jimenez et al., 2020; Njuguna et al., 2020; Pringle et al., 2022; Tompkins et al., 2021; Wadhwa et al., 2021; Wallace et al., 2021; Zawitz et al., 2021). The studies evaluated different testing strategies. Three studied the use of one-time facility-wide testing (Hagan et al., 2020; Pringle et al., 2022; Tompkins et al., 2021), three examined the use of serial or multi-phase testing (Njuguna et al., 2020; Wadhwa et al., 2021; Wallace et al., 2021), two analyzed symptomatic-only testing policies that were later expanded to include some asymptomatic testing (Chan et al., 2021; Zawitz et al., 2021), one compared testing rates across facilities (Chan et al., 2021), and one examined hot spot testing (Epting et al., 2021). All ten were conducted in the spring or summer of 2020 (Chan et al., 2021; Epting et al., 2021; Hagan et al., 2020; Jimenez et al., 2020; Njuguna et al., 2020; Pringle et al., 2022; Tompkins et al., 2021; Wadhwa et al., 2021; Wallace et al., 2021; Zawitz et al., 2021).

### Mitigation Strategy 3: De-Densifications

Five results examined the impact of de-densification on infection rates (Chin, Ryckman, et al., 2021; Jimenez et al., 2020; Leibowitz et al., 2021; Liu et al., 2022; Vest et al., 2021). Vest et al. (Vest et al., 2021), Leibowitz et al. (Leibowitz et al., 2021), and Jimenez et al. (Jimenez et al., 2020) investigated links between how full a prison is and infection rates. Using latent profile analysis, Vest et al. (Vest et al., 2021) found that prisons in Texas that were most effective at reducing outbreaks and deaths operated at 85% capacity or less. Leibowitz et al. (Leibowitz et al., 2021) showed that each 10-percentage-point increase in crowding resulted in a 14% increase in rates of infection (CI, 1.03-1.27) in 14 Massachusetts state prisons. Jimenez et al. (Jimenez et al., 2020) found that among different detention facilities in Massachusetts, the incidence rate was higher in facilities that released a smaller proportion of their baseline population (Jimenez et al., 2020). All three studies concluded that decarceration may be an effective mitigation measure. Chin et al. (Chin, Ryckman, et al., 2021), Leibowitz et al. (Leibowitz et al., 2021), and Liu et al. (Liu et al., 2022) demonstrated that having a smaller number of cellmates reduces the risk of COVID-19 infection for people who are detained in state prisons. Chin et al. (Chin, Ryckman, et al., 2021) found that dormitory residents had higher infection rates than cell residents (adjusted hazard ratio, 2.51; 95% CI, 2.25-2.80). Leibowitz et al. (Leibowitz et al., 2021) found that for each 10-percentage-point increase in single cell units in facilities, the incidence rate decreased by 18% (95% CI, 0.73-0.93). Liu et al. (Liu et al., 2022) used cross-sectional antibody testing to show that having eight or more cellmates was associated with an increased likelihood of previous infection relative to zero or one cellmate (OR, 1.8; CI, 1.0-3.3). All three studies used data from 2020 and early 2021.

### Mitigation Strategy 4: Limited Movement

Three papers, Dunne et al, (Dunne et al., 2021) Brinkley-Rubinstein et al. (Brinkley-Rubinstein, LeMasters, et al., 2021), and Chin et al. (Chin, Ryckman, et al., 2021), used retrospective analyses of longitudinal data to link COVID-19 prevalence to limiting unnecessary contact with other people. Dunne et al. (Dunne et al., 2021) linked two outbreaks in Idaho correctional facilities with work-release programs at food processing plants. They established that correctional facilities with work-release programs should implement measures to mitigate the transmission of COVID-19. Brinkley-Rubinstein et al. (Brinkley-Rubinstein, LeMasters, et al., 2021) found that there was a positive correlation between prison transfers and COVID-19 prevalence three to five weeks after transfers took place. Consequently, they concluded that limiting transfers is an effective measure to reduce COVID-19 transmission. Chin et al. (Chin, Ryckman, et al., 2021) applied cox proportional hazard models to infection data from California state prisons to show that residents of rooms with labor participation had higher rates of infection than residents of other rooms (adjusted hazard ratio, 1.56; 95% CI, 1.39-1.74).

### Mitigation Strategy 5: Masking

Using antibody testing and a questionnaire administered to jail residents, Liu et al. (Liu et al., 2022) investigated contributors to COVID-19 infection in four California jails. They found that having access to new masks more frequently (once a week compared with more infrequently than that) was associated with a decreased risk of infection (OR 13.8; 95% CI 1.8-107.0) (Liu et al., 2022).

### Mitigation Strategy 6: Contact Tracing

Unruh et al. (Unruh et al., 2021) analyzed the use of contact tracing informed by video monitoring in a large juvenile detention center. They found that contacting tracing, which prompted CDC isolation of quarantine guidelines, was beneficial and cost-effective because no secondary cases (i.e., people who were contact traced) led to additional infections.

## Discussion

Our results identify testing; decarceration; limiting transfers or out of room work; improving access to masks; and contact tracing as effective strategies to mitigate the spread of COVID-19 and influenza in jails, prisons, and other detention facilities. Collectively, the scope of research on these topics provides a preliminary scientific understanding of how to respond to respiratory infectious diseases in these settings.

Although the quantity of literature on COVID-19 in prison offers hope that the spread of infectious diseases in jails and prisons will continue to receive more attention, the dearth of literature on influenza should serve as a warning of how easily incarcerated and detained people are forgotten when an emergency no longer accentuates the dire nature of infectious diseases in detention facilities. Given similar pathways of transmission between the two viruses, COVID-19 mitigation interventions in jails and prisons can likely be applied to stop the spread of influenza and other respiratory viruses with similar positive results (Huang et al., 2021; Solomon et al., 2020; Youssef et al., 2022).

Despite the ever-increasing threat of emergent respiratory pathogens (Carlson et al., 2022; Vargas-Parada, 2022), only one result evaluated a proactive pandemic preparedness instead of reactive pandemic response intervention (Lee et al., 2014). The neglect of pandemic preparedness in the literature aligns with overarching under-preparedness of individual facilities across the United States. Few detention facilities have the necessary resources or public health expertise to independently respond to emergent pathogens (Widra & Wagner, 2020). Overwhelmingly, facilities lack pandemic response protocol, despite evidence supporting them (Lee et al., 2014). By comparison, 70% of school districts and an estimated significant majority of nursing homes had plans for how to respond to infectious diseases in early 2020 (“Are Schools Prepared to Respond to a Pandemic?,” 2020; Jones et al., 2020). Unpreparedness undoubtedly contributed to the U.S. government’s ineffective response to the pandemic in jails and prisons during the early stages of the COVID-19 pandemic (Kwan et al., 2022; Maani & Galea, 2020; Widra & Wagner, 2020).

To avoid similarly tragic outcomes in the future, all detention facilities in the United States should develop pandemic response protocols informed by the results of this study. In doing so, questions around how to de-densify facilities; where to quarantine exposed and infected individuals; how to acquire necessary supplies; how to provide medical care; and how to maintain acceptable living standards should be addressed by protocol before outbreaks happen so that facilities are prepared to manage them. Specifically, we suggest developing a protocol for applying the effective mitigation strategies that emerged in this review, loosely grouped into population control, institutional/structural changes, testing, personal protective equipment, treatment, and vaccination. Population control includes plans for decarceration and modifications to movement throughout the facility. Institutional/structural changes include ventilation, improved cleanliness, and access to soap and detergents. Testing includes access to supplies and laboratories for testing. Personal protective equipment includes gloves, masks, and gowns. Increased federal, state, and local financial support towards operationalizing pandemic response committees is necessary, including devoting time for people working in jails and people working in the public health sector to develop, maintain and update protocols.

Results show collaboration between public health and corrections professionals, as 17 out of 28 results had a co-author affiliated with a corrections-based organization. Of note, only 6 out of 28 studies had a corrections affiliate as a first or last author for reference. People who work in corrections may not have the time to support initiatives to improve infectious diseases care or may feel like it is not part of their job. We believe that as the people working in jails and prisons, they have the most experience to lead the development of feasible, practical, and efficient systems of infection control. However, they must receive support from other cross-sector stakeholders. No studies investigated or reported the importance of collaboration between cross-sector stakeholders in infectious disease mitigation. Only nine states have well-developed relationships between the state’s department of health and departments of corrections (Hamblett et al., 2022), and people who are detained are often neglected by pandemic preparedness planning at the local, state, and federal levels of government (Amaning, 2020; Council, 2022). Investment in building sustainable relationships between local, state, and federal public health experts with people who work in carceral systems is crucial. Historically, the decentralized nature of the U.S. criminal justice system has been a barrier to building public health-carceral partnerships, as it consists of several systems (jails, prisons, parole, etc.) overseen by different authorities. Robust and active relationships between stakeholders are essential to assist individual facilities, which often lack essential public health expertise and resources, effectively prepare for and respond to pandemics (Hamblett et al., 2022; Puglisi et al., 2022). One way to facilitate this is to have facility-level pandemic preparedness committees with diverse representation from cross-sector representatives. These committees should meet monthly to develop and update preparedness protocol. Identifying champions at detention facilities willing to lead pandemic response preparation is also propitious.

Finally, results largely focused on the impact of interventions on incidence or mortality. In doing so, they failed to evaluate or acknowledge the measures’ impact on quality of life. For example, few results considered the implications of mitigation measures on mental health, despite links between multiple interventions we identified and negative mental health outcomes. Moving forward, research should contextualize interventions within a larger set of priorities to help facilities and policymakers holistically evaluate the impact of mitigation measures, so they can maintain an acceptable standard of living for people who are detained in the event of an outbreak.

One limitation of our review is that we only used the PubMed/MEDLINE and Embase databases. This review, as is true generally with scoping reviews(Arksey & O’Malley, 2005; Peters et al., 2015; Pham et al., 2014), does not assess the quality of publications. Consequently, it should only be used to assess what evidence exists on this topic, not if that evidence is robust and generalizable or if there are gaps in the literature from low-quality research. This is particularly true because not all results included in this study were peer-reviewed. Another limitation is that through requiring all results to report real-world data on the impact of an intervention, some results were excluded that may be relevant to the topic at hand. This was done to exclude results editorializing on interventions without scientific backing. However, this also excluded studies such as cost-benefit analyses and modeling studies that may be informative.

## Conclusion

This scoping review aggregated studies on the effectiveness of influenza and COVID-19 infection mitigation strategies in detention facilities. It shows that vaccination; testing; decarceration; limiting transfers or out-of-room work; improving access to masks; and contact tracing are effective measures for mitigating the spread of influenza and COVID-19 in these settings. Using these strategies as keystones, all U.S. detention facilities should develop a pandemic preparedness protocol to keep people safe when the next pandemic arises.

## Data Availability

All data produced in the present study are available upon reasonable request to the authors

## Acknowledgements

This research was supported in part by the Tupper Research Fund at Tufts Medical center. We would like to thank Tufts University School of Medicine library staff for their guidance in determining search terms.

## Author Confirmation Statement

All authors contributed significantly to the manuscript. LKW and YN independently screened search results and extracted data from search articles. Emily assisted with drafting and editing the manuscript. AGW and MT came up with the idea for the project and oversaw the data collection, analysis and writing of the manuscript.

