## Appendix 1 for "COVID-19 and Influenza Infection Prevention and Control Measures in U.S. Jails, Prisons, and Detention Centers: A Scoping Review"

Appendix 1: PubMed Search Strategy

((prison* OR jail* OR carceral* OR detention* OR detain* OR incarcerate* OR correctional OR penal) AND (COVID-19 OR SARS-CoV-2 OR influenza OR flu)) AND (1982:2022[pdat])) AND ("United States"[Mesh] or "united states") AND ((humans[Filter]) AND (english[Filter]))
